# Study Protocol for a Research and Development Project: Optimizing a Unified Parent Training Intervention to Prevent Child Mental Health Problems and Neglect

**DOI:** 10.1101/2022.08.16.22278822

**Authors:** Truls Tømmerås, Agathe Backer-Grøndahl, Anne Arnesen, Anett Apeland, Hanne Laland, Elisabeth Askeland, John Kjøbli, Thormod Idsøe, Anette Arnesen Grønlie, Maria Begõna Gomez, Line Ragna Aakre Karlsson, Elise Dyrkoren, Sissel Torsvik, Andreas Høstmælingen, Kristine Amlund-Hagen, Marion Forgatch, Phillip Andrew Fisher

## Abstract

This protocol describes a research and development (R&D) project aimed at optimizing a targeted, preventive, efficacious, and tailored intervention called Supportive Parents – Coping Kids (SPARCK). Combining recent developments in basic and intervention research, the goal of this project is to develop, test and optimize a unified parent training intervention targeting children who display externalizing and internalizing symptoms and parents who are at risk of exhibiting maladaptive or neglectful parenting behaviors. We plan to utilize various design-based research methodologies to investigate what works for whom in which context, points which are essential to the innovation process, by employing a mixed methods research design and an iterative optimization process of testing and refinement. Furthermore, we introduce a cocreation process for SPARCK to involve relevant stakeholders working in Norwegian frontline services for children and their families to ensure that the intervention adheres to the needs and constraints encountered by these stakeholders and thereby promote the scalability and sustainable implementation of SPARCK. In this paper, we present the theoretical and methodological background of this approach to R&D in the field of mental health prevention as well as the operationalization of innovative methodology in the current project. This R&D approach aims to produce new knowledge concerning individual change mechanisms in parent training interventions and stakeholder feedback pertaining to intervention components and implementation strategies, all of which are imperative for the iterative SPARCK design process.

## Introduction

More than 10 percent of Norwegian experience mental health problems (Reneflot et al., 2018). These young people are at risk of negative developmental trajectories, including decreased quality of life, poor mental and somatic health, social and academic difficulties, school absenteeism and drop-out, and subsequent exclusion from social and working life (Caspi et al., 2016; Gubbels, van der Put, & Assink, 2019; Nordmo et al., 2022). It is imperative to prevent such negative life-course trajectories. However, some evidence suggests that preventive frontline services lack effective interventions to prevent mental health problems. The quality of intervention in frontline services for children and families varies tremendously, and this topic has been highlighted as one of the main challenges for municipal frontline services, such as Child Welfare Services (CWS), Education and School-Psychology (PPT) services, and mental health services (Auditor General, 2021). Accordingly, there is a high demand for effective interventions in frontline services.

This project builds on two decades of development and evaluation of the Norwegian versions of the Parent Management Training Oregon Model (PMTO) and specifically the Norwegian short-version PMTO derivate, Brief Parent Training (BPT), which targets children with externalizing problems (conduct and oppositional problems; Forgatch & Kjøbli, 2016; Forgatch & Patterson, 2010; Solholm, Kjøbli, & Christiansen, 2013). Supportive Parents – Coping Kids (SPARCK) intervention incorporates new components to broaden the target group of parent training to include children displaying internalizing symptoms (anxiety and depressive symptoms) and/or caretakers experiencing challenges with child rearing (those who are at risk of maladaptive or neglectful parenting). Methodologically, the project is inspired by challenges and recent developments in evaluation and implementation science. In that regard, the project incorporates a design-based methodology mainly drawn from the IDEAS impact framework (Schindler, Fisher, & Shonkoff, 2017) and establishes a long-term plan for innovation of a parent training intervention to suit the needs of Norwegian frontline services. This protocol presents the background and the design-based methodology of this project, including a well-specified theory of change (ToC), cocreation in research and development, and a mixed methods research design intended to produce knowledge that is useful for the design process.

### Needs for Innovation

Despite several decades of development and evaluation of evidence-based interventions (EBIs) to promote positive child development, several interrelated challenges concerning EBIs remain. EBIs continue to exhibit small to moderate effect sizes (Duncan & Magnuson, 2013; Supplee & Duggan, 2019; Weisz et al., 2017), and few children and families have access to interventions that have been documented as effective (Christiansen, 2015; Glasgow et al., 2012; Nøkleby, Johansen, Jardim, & Muller, 2020; Skogen & Torvik, 2013). Thus, the public health impact of EBIs is probably low. Moreover, the nature of mental health symptoms often transcends traditional symptom domains, and comorbidity of mental health problems in children is very common (Caspi et al., 2020). Children often display both internalizing and externalizing symptoms (Caspi et al., 2020; Marchette & Weisz, 2017). The traditional assumption that the identification of one mental health diagnosis or problem identifies the root cause of the problem and thus indicates the manner in which treatment should be delivered is a poor match with the comorbid and transdiagnostic realities experienced in the context of regular practice. This difficulty limits the ecological validity of standard EBI protocols, which are often developed to address one diagnosis or problem domain (Lyon, Dopp, Brewer, Kientz, & Munson, 2020). Adding to this complexity, there is a large degree of heterogeneity in frontline services in terms of professional level and clinical expertise as well as the resources that are committed to client contact (Tommeraas & Ogden, 2015). Accordingly, EBI protocols should be designed in light of this heterogeneity among clients and service systems as well as the need for flexible, efficient and usable interventions. The lack of ecological validity in EBIs may have contributed to the belief that these interventions are challenging to implement and sustain in regular practice settings and thus contributed to the gap between science and practitice as well as the limited scope of EBIs (Lyon et al., 2020).

A limited methodological scope in the context of evaluation may represent another challenge for innovation in the context of EBIs. The predominant methodology in EBI and evaluation science has been to evaluate EBI program packages using randomized controlled trials (RCT; Collins, 2018; Lyon et al., 2020; Schindler, McCoy, Fisher, & Shonkoff, 2019). Program package evaluation in RCT designs via the display of program package group mean effects has been vital for the progress of the EBI field. However, RCTs mask within-group heterogeneity in terms of intervention effects and active intervention ingredients, a difficulty which is often referred to as the black box problem in the context of EBIs (Kazdin, 2018; Lyon & Koerner, 2016; Schindler et al., 2019). The race to attain a high degree of internal validity in the context of expensive and time-consuming RCTs must be complemented with a research focus that allows us to look inside this black box and identify treatment nonresponders and the components that are effective for individual users; all of this information is necessary for innovation in the context of EBIs (Supplee & Duggan, 2019). The black box problem and the limited scope of EBIs warrant a shift in focus to the evaluation methodology that is useful for the design process alongside the more traditional and sorely needed evaluation of treatment packages in RCTs. The inclusion of various design-based methodologies to produce knowledge concerning the active ingredients of interventions may accelerate innovation and ultimately answer the following question: What works for whom in which contexts? (Collins, 2018; Schindler et al., 2017; Supplee & Duggan, 2019).

### Design-Based Methodology

The need for intervention innovation is reflected in the necessity of additional design-based methodological elements that can be useful for the innovation process. First, a clear and precise specification of a Theory of Change (ToC; also known as a causal model or conceptual model) is a prerequisite and a tool for a rigorous research and development (R&D) process. A ToC refers to “a detailed set of beliefs about observable changes that are expected from an intervention” (p.147; Schindler et al., 2019). A well-specified ToC is vital to improve precision in the development and testing of an intervention (Collins, 2018; Supplee & Duggan, 2019). It is a tool for understanding what components and procedures work, why they work, for whom they work, under which circumstances they work, and on what outcomes they work. A well-specified and conceptualized ToC may serve as a tool for operationalizing the findings of basic and intervention research to advance the research-based development of EBIs. Moreover, a clear and conceptualized ToC is vital to the establishment of common ground in a design process featuring cocreation (Schindler et al., 2019).

Second, incorporating knowledge drawn from relevant stakeholders into the R&D intervention is pivotal for an effective design process (Klev & Levin, 2016). Hence, cocreation featuring relevant stakeholders (i.e., client families, service counselors and leaders) as well as program developers and researchers, both with respect to the research and the development of the intervention, will likely influence the usability and ecological validity of the EBIs (Lyon et al., 2020), allowing EBIs to be designed to fit stakeholder needs, knowledge, and service constraints. Ultimately, the cocreation of interventions will likely impact the scalability and sustainability of EBIs (Lyon & Bruns, 2019; Mummah, Robinson, King, Gardner, & Sutton, 2016).

Third, compared to traditional RCT designs, alternative evaluation designs are more appropriate to the task of addressing the active components and change mechanisms involved in the intervention. For example, the alternatives include sequential multiple assignment randomized trials, factorial experiments, single case experiments, or mixed methods designs (Collins, 2018; Kazdin, 2018; Palinkas, Horwitz, Chamberlain, Hurlburt, & Landsverk, 2011; Supplee & Duggan, 2019). Particularly, the latter two designs are well suited to rapid-cycle and iterative testing, in which context research hypotheses can be subsequently raised and tested, with the aim of moving rapidly toward higher levels of evidence, which is essential during the early stages of a design process (Gallo, Comer, & Barlow, 2013; Supplee & Duggan, 2019).

Innovative design-based frameworks that can accelerate EBI progress are emerging. These frameworks often share an introspective focus on active components as tools for innovation, and they propose different methodological elements that are important to the design process (Collins, 2018; Lyon et al., 2020; Schindler et al., 2017). One such model for the science-based development of interventions is the Frontiers of Innovations’ IDEAS Impact Framework (Innovate Evaluate Adapt Scale; Shonkoff et al., 2016). IDEAS highlights several methodological elements, such as cocreation in the context of intervention research and development to reflect stakeholders’ needs; precision in ToC specification and assessment to understand what works for whom and how; rapid-cycle iterative testing and optimization to accelerate our ability to gather evidence; and the establishment of an evaluation plan combining research, theory, and practice via the optimization of a ToC and the intervention materials (Schindler et al., 2017).

### The Current Project: Aims and Research Questions

Incorporating a design-based methodology mainly based on the IDEAS impact framework but also on the multiple optimization strategy (Collins, 2018) and a user-centered design (Lyon & Koerner, 2016), the goals of this project are to (1) broaden the scope of the BPT-based intervention to reach a potentially large group of children and families in need of targeted (selected and indicated) preventive frontline services; (2) to design and optimize a ToC based on the BPT/PMTO and new knowledge drawn from relevant basic and intervention research; and (3) to cocreate a flexible and usable intervention that is tailored to stakeholders’ knowledge and needs. Thus, the R&D pertaining to the new intervention SPARCK is based on the constraints for the intervention’s development, including societal needs for mental health intervention, service constraints such as resources and personnel, and client needs for individualized and tailored intervention, with the aim of creating a scalable and sustainable intervention for routine practice.

We will test individual effects using single case experimental designs (SCEDs), systematic stakeholder feedback collected via qualitative interviews, and biomarker observations referencing parent and child cortisol stress hormones collected from hair samples. With the participation of service counselors and clients, we will cocreate a SPARCK prototype that combines BPT and new knowledge drawn from basic and intervention research concerning comorbid mental health symptoms and active ingredients for the target group, respectively. The aim of this effort is to design a unified and transdiagnostic preventive intervention with a broad theoretical and empirical foundation to address the heterogeneity in the challenges encountered in regular practice settings. All aspects of the SPARCK prototype, such as the intervention components, sequencing, dosage and program materials, will be optimized throughout the development and research phases, with the aim of achieving higher levels of efficacy and usability in the complete version of SPARCK. In addition, we will explore and cocreate implementation strategies that may further promote SPARCK’s scalability and sustainability in frontline settings.

The following research questions serve as a starting point for the R&D process.

- What families/children seem to benefit or not benefit from the SPARCK intervention?
- What seem to be the active components and change mechanisms for individual families?
- Is the ToC and the accompanying program material included in SPARCK usable and feasible for counselors working in Norwegian frontline services?
- What is the appropriate structure and sequencing of procedures and components for different users of SPARCK (including dosage, fixed component order vs. customized adaptation, decision support, etc.)?
- What implementation strategies are feasible and appropriate for supporting the sustainable implementation of SPARCK in the context of Norwegian frontline services?

The project has the potential to contribute new knowledge to the mental health prevention field in general, with respect to the benefit of utilizing innovative methodology in the intervention design process, as well as evidence concerning both the change mechanisms underlying a unified parenting intervention to promote healthy child development and the selection and tailoring of implementation strategies.

## Methods

The innovative research methodology used in the R&D design process is displayed in Figure 1. The following core activities serve as the basis for the development phase and the empirical testing of SPARCK in the subsequent research phase: 1) identify and define the constraints for development; 2) search the literature and develop a prototype ToC, including the development of an intervention manual and materials; 3) establish the research design and assessment strategies in accordance with the project aims and the available resources; and 4) recruit and train a lab of service counselors.

**Figure 1.**
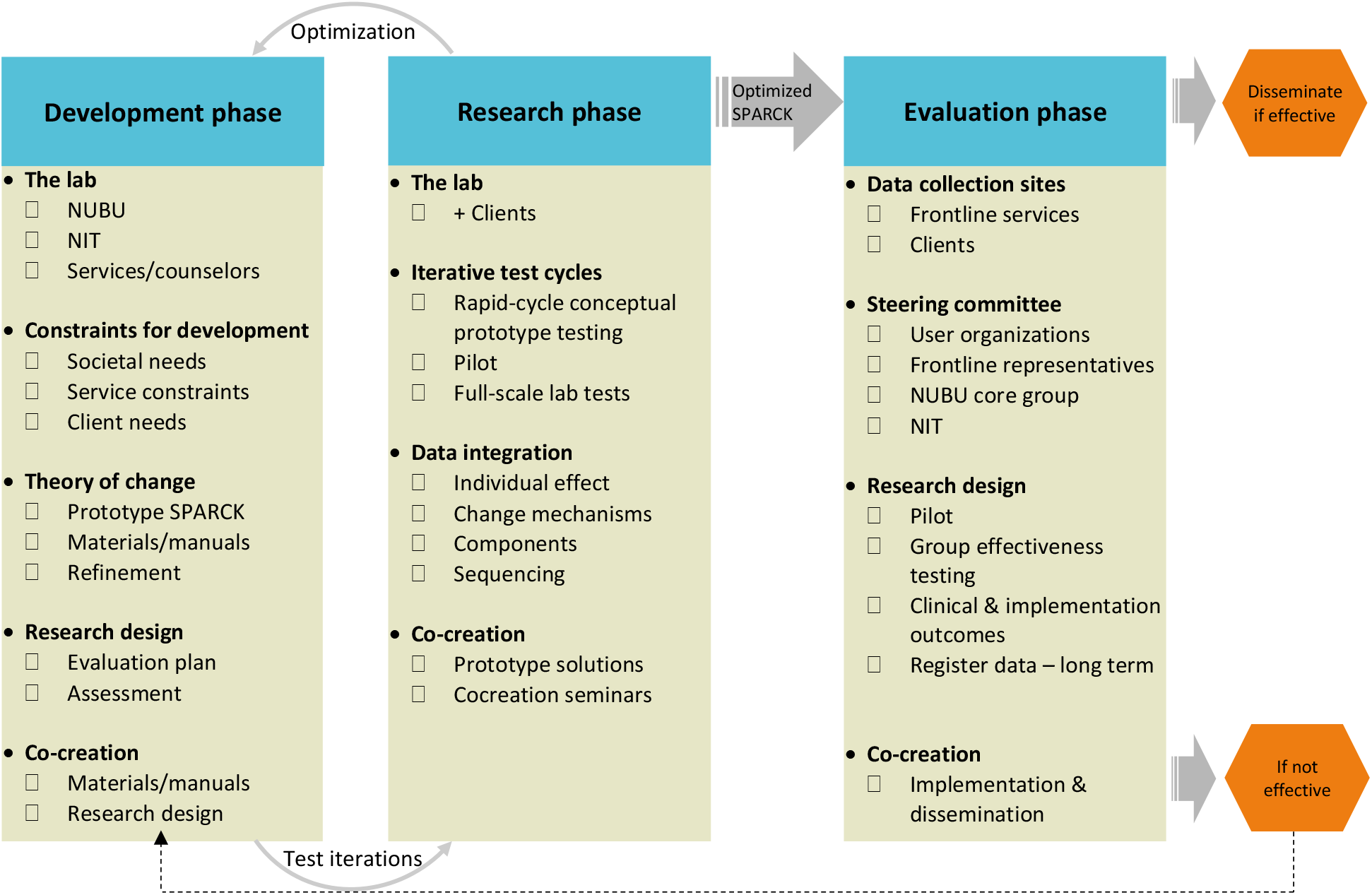
Evaluation plan for SPARCK

In the research phase, the core activities concern the following tasks: 1) recruitment of client participants; 2) execution of iterative test cycles, including rapid-cycle conceptual prototype testing and refinement of the conceptual version of ToC (Chen, Neta, & Roberts, 2021), pilot testing SPARCK, and full-scale test cycles in frontline services; 3) a period of data analysis and the elaboration of prototype optimization solutions; and 4) a research phase including cocreation activities such as lab seminars and meetings to present and receive feedback concerning the results and the prototype optimization solutions.

This iterative design process involves shifting back and forth between the development and research phases so that an optimized version of the ToC for SPARCK can be tested in a subsequent cycle. The optimization of a usable intervention that is well suited to family and service constraints implies a process of refining and adding new content and materials, as well as, which is equally important, the removal of redundant content to increase precision and reduce complexity in SPARCK.

### Developing SPARCK: Prototype Theory of Change

There is strong evidence to suggest that parent–child interaction is a mediator that is associated with long-term outcomes for children, including maltreatment (Green et al., 2018), internalizing problems (Shimshoni, Shrinivasa, Cherian, & Lebowitz, 2019), and externalizing problems (Nærde, Janson & Stoolmiller, in review). As a result, a parenting intervention in which parents are the agents of change is of particular relevance to a unified preventive intervention during childhood. In the initial design of the prototype ToC in SPARCK, we will follow the following tenets: 1) integrate new and relevant evidence drawn from basic research; 2) develop the model based on different theoretical intervention models to promote flexible ways of understanding and tailoring interventions to family needs; 3) incorporate new evidence from intervention research and identify the active components contained in effective interventions; 4) search for transdiagnostic approaches and components that may reduce the complexity of interventions and broadly promote healthy child development.

The prototype ToC is displayed in Figure 2. To adapt and tailor the intervention content and to promote different perspectives on ways of addressing individual family needs, the ToC of SPARCK is to some extent a theoretical hybrid, and the intervention components are influenced by different causal theories.

**Figure 2.**
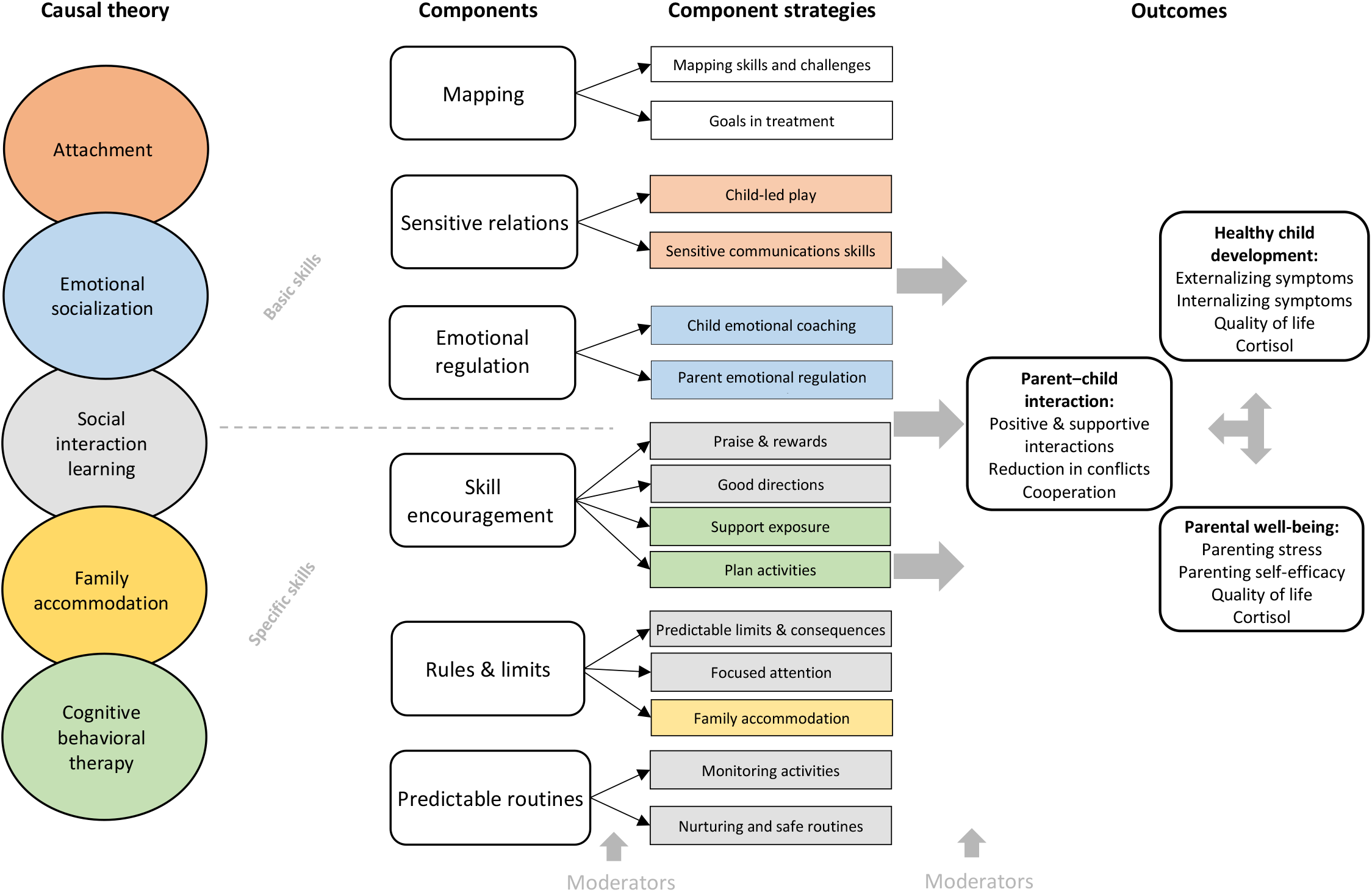
SPARCK theory of changes in conceptual prototype

First, the causal theory included in BPT/PMTO, i.e., social interaction learning theory (SIL; Patterson, 2016), and the BPT component strategies that target externalizing problems serve as a point of departure for SPARCK (Forgatch & Patterson, 2010; Solholm et al., 2013). SIL highlights the way in which externalizing problems develop in the context of coercive parent–child interactions and the reciprocal process of negative reinforcement between children’s maladaptive behaviors and parents’ withdrawal and suboptimal parenting behaviors (Patterson, 2016).

Second, insecure attachment, particularly disorganized attachment, has been broadly associated with negative short- and long-term outcomes for children, such as poor emotional and self-regulatory capacities, mental health problems, cognitive problems, and dysregulation of cortisol hormones (Bernard et al., 2012; Cyr, Euser, Bakermans-Kranenburg, & Van Ijzendoorn, 2010; Euser, Alink, Stoltenborgh, Bakermans-Kranenburg, & van IJzendoorn, 2015). Parenting capacities constitute a core aspect of secure attachment (Cyr & Alink, 2017), and the literature concerning sensitive and nurturing parenting practices has influenced the ToC of SPARCK.

Third, emotional dysregulation has been associated with disrupted parenting practices as well as internalizing and externalizing symptoms in children (Fraire & Ollendick, 2013; Morris, Criss, Silk, & Houltberg, 2017). SPARCK is influenced by emotion socialization theory, which emphasizes the ways in which parents themselves can become aware of and learn to regulate their own emotions (Gottman, Katz, & Hooven, 1996) as well as the ways in which they can help their children deal with their emotions via emotional coaching (Johnson, Hawes, Eisenberg, Kohlhoff, & Dudeney, 2017). Emotional socialization interventions have been found to prevent and reduce mental health problems in children (Ansar et al., 2022; Bølstad et al., 2021; Edrissi, Havighurst, Aghebati, Habibi, & Arani, 2019).

Fourth, it has been suggested that parents could also be a mediator of change in interventions targeting internalizing symptoms. Support for parent/family-based cognitive behavioral therapy (CBT) has been reported by systematic reviews focusing on the treatment and prevention of anxiety disorders in young children (Comer, Hong, Poznanski, Silva, & Wilson, 2019; Schwartz, Barican, Yung, Zheng, & Waddell, 2019). SPARCK is influenced by the behavioral components of CBT by helping parents with respect to exposure to fears and behavioral activation to promote coping and specific skills that are necessary in daily life.

Fifth, the concept of family accommodation is incorporated in the ToC of SPARCK. Family accommodation refers to change parents make to their behavior, such as facilitating avoidance, excessive support and modifying family routines, to help the child mitigate feelings of distress and anxiety, which can negatively reinforce children’s problems and contribute to the maintenance or worsening of symptoms (Lebowitz, Panza, & Bloch, 2016). Compelling findings have shown that parental accommodation plays a significant role in the maintenance of anxiety symptoms (Shimshoni et al., 2019).

Even though SPARCK is inspired by different causal theories and components, there is some degree of overlap among these influences. For instance, several theories highlight the need for sensitive and responsive interactions. Such components that are common across theories of parent counseling may represent common components that are assumed to be important across a variety of problems and diagnoses. Other components are specific to one theory and address the specific problems for which the theory was originally developed. Thus, SPARCK is constructed based on the idea of utilizing general common components in combination with specific components that are tailored to specific problems. Hence, SPARCK consists of parenting components that are considered to be important for supporting children and families who display a broad range of problems. The conceptual prototype ToC, which is displayed in Figure 2, serves as a foundation for operationalizing component strategies in the SPARCK manual and intervention materials as well as a guiding tool for research focusing on mechanisms of change or the lack thereof.

A critical consideration in this context is the sequencing of the components and target strategies. Ultimately, two possibilities must be considered in the context of SPARCK: fixed sequencing versus customized adaptive sequencing. Ease of use may suggest linear sequencing; in contrast, adaptive sequencing may have an impact on the complexity of delivery, but it also allows for more flexibility to individualize content. In the SPARCK prototype, we start with a mixed solution featuring a linear basic sequence and an adaptive sequence, according to which all families will receive components listed above the dotted line in Figure 2 before the customization of the components listed below the dotted line. This solution may balance the need for personalized and individual tailoring and dosage with ease of use in the context of frontline services.

### Recruitment of Service Counselors and Clients

An important aspect in this project is *the lab*, which includes stakeholders, professional counselors, their leaders, and end user families recruited from frontline services. The professional counselors will include 14 parent counselors who are accustomed to working with EBIs; these counselors will be recruited from seven municipalities that are representative of all Norwegian health regions. The **counselors** are recruited based on the following criteria: (a) they are trained in PMTO, (b) they are motivated to participate in the project, (c) they have access to the target groups, (d) they can deliver cases to research as part of their regular practice, and (e), they represent the relevant frontline service sectors that provide interventions to children and families. These counselors will represent municipal frontline services, comprising CWS services, PPT school-psychology services, and health services, all of which deliver some sort of prevention measures and/or treatments to children, youths, and families who struggle with socioemotional problems and parent–child interactions. These 14 counselors will be closely involved in the iterative R&D process as described below. In the following, when we use the term lab, we refer to these counselors.

The inclusion of study participants and client families in the project will follow the formal and clinical screening procedures used by the lab counselors ‘ in their regular frontline services. In frontline services, no diagnosis is necessary to receive help, which reflects the selected and indicated level of prevention. In addition, formal screening procedures will be implemented prior to inclusion in the iterative test cycles (as presented in the assessment procedures). Client participants will typically be parents of children who display internalizing and/or externalizing symptoms and/or parents who are at risk of exhibiting maladaptive or neglectful parenting practices. Clients will be excluded if it is indicated that the target child has been exposed to physical violence or sexual abuse or if children and/or parents are receiving other ongoing treatments related to the target group domains of SPARCK.

### Research and Development Design

#### Cocreation and the iterative process

Cocreation and rapid-cycle iterations are central features of the SPARCK R&D phases. The hub for the R&D activities will be the project’s core group, researchers, clinical psychologists, and research administrative staff working at the Norwegian Center for Child Behavioral Center (NCCBD). Fast-cycle iteration is a process of innovation that includes subsequent testing, learning, and refinement of an intervention, that is, optimization of ToC (Center on the Developing Child, 2022). The R&D process requires quickly incorporating test results into the ToC, and then retesting the optimized ToC in a subsequent cycle. These repetitive cycles of testing generate hypotheses and represent an opportunity to design the SPARCK intervention based on what works or does not work for different families as well as with respect to the possible constraints imposed by the frontline services; thus, it allows us to move toward higher levels of efficacy and usability with respect to SPARCK.

First, the iterations start with feedback collected via the rapid-cycle conceptual prototype testing of the ToC (Chen et al., 2021), in which context the ideas and components displayed in Figure 2 are presented to relevant stakeholders to obtain immediate feedback. The rapid-cycle prototyping will involve the NCCBD core group and the National Implementation Team, which constitute the regional implementation coordinators for the PMTO and BPT in Norway (Askeland, Forgatch, Apeland, Reer, & Grønlie, 2019). The ToC prototype will be optimized alongside the operationalization of component strategies to develop a SPARCK manual. Second, the SPARCK intervention and research design will be pilot tested in a process involving the NCCBD core group and three families recruited from collaborating services in one municipality.

The third stage of the iterative R&D process is that of the “full-scale” prototype test cycles with the lab in collaboration with regular frontline services. User feedback provided by parents, lab counselors, and municipal leaders will be used to optimize SPARCK. In the test cycles as well as in the rapid-cycle prototyping, the cocreation process will feature the NCCBD core group as a hub for the collaborative process. The core group will analyze data and feedback and develop potential optimization solutions. These solutions will be returned to the lab in cocreation seminars, in which the lab will provide advice on the proposed optimization solutions. Accordingly, these lab counselors are of particular importance for the optimization process due to the fact that they are experts on the frontline service system and its clients.

Feedback concerning the SPARCK ToC emerging from the lab will be systematically collected in the test cycles using qualitative and quantitative data (as presented below). Following each test cycle, a period of analysis, cocreation and optimization will ensue, after which the redesigned version will be tested in a subsequent test cycle. Depending on the results, we opt for a maximum of three test cycles. This iterative process implies the need to utilize a research design that allows for small-scale testing and innovation. Therefore, we choose to employ SCED, which is a robust research design for testing individual effects and is ideally suited to fast-cycle testing (Gallo et al., 2013).

##### Mixed methods approach

We use a convergent parallel mixed methods design (Creswell & Creswell, 2017), which combines quantitative SCEDs, biological observation measures, and qualitative semi-structured interview data across multiple respondents. Qualitative and quantitative data will be combined to improve our understanding of whether and how change occurs within each family and each frontline setting.

##### Single case experimental design

SCEDs are assumed to be particularly suitable for testing individual (case) intervention effects (Kazdin, 2018) and for research conducted during the early phases of the testing and development of interventions (Gallo et al., 2013). Small-scale pre-post studies focusing on a single group are frequently conducted as pilot studies for large-scale randomized control treatment studies (Gallo et al., 2013). Such pre-post pilot studies are important to obtain knowledge concerning feasibility and user satisfaction but are less suited to the task of detecting functional relations between independent and dependent variables. In contrast, well-designed (and randomized) SCEDs are assumed to have internal validity equivalent to that of RCTs, thereby facilitating the identification of causal relations (Kazdin, 2018; Kratochwill & Levin, 2014). Instead of comparing group mean level differences between an intervention and a control, in an SCED, we can compare individuals in a baseline (A) phase (i.e., a phase without treatment) with themselves in an intervention (B) phase (Barlow, Nock, & Hersen, 2009). Additionally, SCEDs are superior to regular group designs in that they permit the investigation of the change in target outcomes for each individual, both when introducing the intervention and throughout the B phase (Smith, 2012). Nevertheless, SCEDs can be resource-intensive due to the need for extensive measurement across multiple data points. Adding to these challenges, SCEDs do not profit from “the magic” of large numbers in the normality distribution of data in group designs and thus may be particularly vulnerable to measurement error.

We will employ a nonconcurrent randomized multiple baseline design with replications across participants (Barlow et al., 2009). Randomized phase start-point designs have been suggested to ensure strong internal validity, and replication of participants across frontline settings would add to the external validity (Kratochwill & Levin, 2014). This design is a particular variant of a simple AB design. Participants will be assigned randomly to different lengths of the baseline (A) phase, i.e., to 4, 5, or 6 weeks of baseline, prior to the intervention B phase (lasting for up to 12 weeks). Ideally, we would follow through on baseline assessments until the stability of phenomenon under scrutiny is ensured; however, there are ethical considerations that argue against delaying intervention for the target group in the SPARCK project.

#### Measurements

##### Quantitative measurements

In the first cycle, we will use four different modes of measurement: (1) periodic assessments of parent-reported outcomes across three time points; (2) weekly assessments in the A and B phases; (3) calculation of the average level of cortisol stress hormones in AB phases; and (4) a weekly report by lab counselors concerning SPARCK fidelity and client responsiveness during sessions.

##### Periodic assessments

Case profiles and movement into or out of the clinical ranges of symptoms between, before, and after interventions will be assessed before, midway through, and after interventions in the test cycles. Children’s symptoms will be assessed using the parent-reported *Strengths and Difficulties Questionnaire* (SDQ; Goodman, Lamping, & Ploubidis, 2010; Kornør & Heyerdahl, 2017), *Revised Children’s Anxiety and Depression Scale* (RCADS; Chorpita, Yim, Moffitt, Umemoto, & Francis, 2000), and *Eyberg Child Behavior Inventory* (ECBI; Eyberg & Ross, 1978). Other variables to be assessed include family demographic variables, parent mental distress, and parent self-reported stress.

##### Weekly assessments

In cycle 1, we will explore the assessment of parenting-reported behaviors in a manner that is exclusively constructed to represent each component strategy in the SPARCK ToC (see Figure 2). Examples include “I help my child put their feelings into words” and “I encourage my child to follow through if he or she is anxious in daily situations”. We have attempted to develop formulations that are generically meaningful to allow for assessment in both the baseline and the intervention phases. Because not all parents employ each SPARCK component strategy, the measurement model for each “case” may be individualized in accordance with the component strategies provided during sessions. Child internalizing and externalizing outcomes will be assessed weekly using the parent-reported *Behavior and feelings survey – caregiver report* (BFS) to measure children’s mental health symptoms (Weisz et al., 2019). The 12-item brief problem checklist contains six items pertaining to the internalizing subscale and six items concerning the externalizing subscale.

##### Biomarker observation

To observe potential biological changes following intervention, we will employ an assessment of the stress hormone cortisol collected from scalp hair samples. Assessment of cortisol deposited in hair provides an opportunity to observe individual changes in average levels of stress hormones without the confounding influences of daily and situational fluctuations that can affect blood, saliva, and urine samples, which are also frequently used (Greff et al., 2018). Hair cortisol samples provide the opportunity to assess changes in average levels of cortisol for 16 weeks per 4 cm sample. Depending on the diameter size of the hair samples, 0.5 segments or 1 cm segments can be analyzed (>0.4 mg per datapoint/segment; (Technische Universitat Dresden, 2020). Thus, up to 8 cortisol data segments can be acquired per sample. We will assess cortisol in parent and child dyads in the baseline and intervention phases. For practical and economic reasons, only one parent, namely, the parent who spends the most time with the child and who completes the parent-reported questionnaires, will provide hair samples.

##### Lab counselors’ weekly questionnaires

A weekly measure of counselor-reported fidelity and client responsiveness will be constructed exclusively for this project. The purpose of this measure is to assess the use of component strategies, pedagogical tools used during sessions, and client responsiveness to the component strategies provided during the sessions. Moreover, the counselors will report parents’ ratings on a three-item and five-point scale, i.e., the *Goals in Intervention* measure, to assess the families’ goal achievements in the SPARCK intervention phase. This rating represents both a clinical strategy in the Mapping and assessment component as well as a quantitative outcome measure during the intervention phase. In a joint process during the first session, i.e., the Mapping component, parents and counselors will set goals for these three items based on several guiding criteria, including the requirement that the goals should target concrete child symptoms/skills, parenting practices or parent–child interactions that are possible to address during SPARCK sessions. These goals will be scored by the parents at the beginning of each session and reported by the counsellor. *The Goals in Intervention* measure is adapted from the Top Problem Assessment Scale (Weisz et al., 2011) to suit the preventive context of SPARCK.

##### Qualitative Measurements

Semi-structured qualitative interviews will be collected posttreatment from families, counselors, and leaders. These interviews will be analyzed using thematic analysis (Braun & Clarke, 2006) prior to being integrated with the quantitative data to examine change mechanisms and experienced benefits and outcomes. In addition, the qualitative data will serve as an instrument for the systematic collection of stakeholder feedback as well as to access contextual barriers and facilitators to suggest implementation strategies that can be used to support SPARCK sustainability in practice (Waltz, Powell, Fernández, Abadie, & Damschroder, 2019).

To varying degrees, the interview guides will address similar thematic content across caretakers, counselors, and leaders. This content includes the feasibility, appropriateness, acceptability and usability of the SPARCK intervention (Lyon & Koerner, 2016; Proctor et al., 2011) and the benefits of the intervention for users with respect to the needs of families and service counselors as well as related advantages and outcomes. Other topics that will be covered include recruitment, training and supervision, program materials, intervention content fit and sequencing, and implementation facilitators and barriers. The interviews will last up to 1.5 hours each. For the leaders, we plan to conduct focus groups featuring the counselors ‘ closest manager and one senior manager. Notes from the lab cocreation meetings and reports from SPARCK supervisors will also be included as qualitative data for the optimization of SPARCK content.

### Results – Optimization Decisions

The number of iterative cycles that should be conducted depends on the results obtained in each cycle. In cycle 1, we plan to test the ToC in the context of a limited age span and symptom domain, including parents and their children, with the latter being between the ages of 6 and 10 years and displaying signs of internalizing problems. The main reason for this approach is to reduce participant heterogeneity in the first cycle to ensure sufficient replications of relatively homogenous cases in terms of age and symptom domain. Moreover, we opt to test all components of ToC in cycle 1 to learn more about their applicability to and efficacy for families. Depending on the results obtained in cycle 1, subsequent cycles will include the remaining age spans and/or symptom domains to test the SPARCK ToC with respect to the complete target group. Moreover, depending on the results and feedback obtained from stakeholders, we may also modify the research design and add and/or remove measurement instruments.

A period of analysis, cocreation activities and optimization will follow each test cycle. The results obtained via the different measures will be combined and synthesized to determine what works or does not work for various subjects and to try to reveal systematic patterns of change or a lack of change both in and between cases. Similarly, we hypothesize that these qualitative data will contribute to improving our understanding of the change mechanisms associated with families in combination with the quantitative measures. In cases featuring discrepancies between measurement instruments and/or methods, qualitative evaluations will inform the optimization decisions. It is important to note that we will not use predefined criteria, such as effect size cutoffs, to determine optimization decisions. The decision concerning when to transition between phases in the innovation plan will be based on data integration and a determination of when the optimization changes are approaching a satisfactory level of detail, indicating that an acceptable level of confidence regarding who will benefit from SPARCK has been achieved.

### Ethical considerations

The project has been reviewed and approved by the Regional Committees for Medical and Health Research Ethics, Southern and Eastern Norway (REK South East). The project involved certain ethical considerations concerning the families receiving SPARCK and their potential vulnerable life situation. However, SPARCK builds on well-established and tested components, indicating that its content is safe and robust. We will use the University of Oslo’s services for sensitive data (TSD) for data management, including collection, handling, analysis and storage procedures and follow standards of collection and storage of biological material. Participants will sign written informed consent forms prior to their inclusion in the project, and their anonymity and confidentiality will be secured in the project’s outputs.

## Concluding Remarks

By utilizing a design-based methodology, the overarching goal of this R&D project is to cocreate and optimize a usable and efficacious intervention focused on a broad and unified target group to promote scalability and sustainability with respect to the regular provision of services. Thus, SPARCK has the potential to reach large user groups in Norwegian frontline services. In the iterative R&D process, we aim to provide empirical evidence that can be useful for innovation regarding SPARCK and to transition toward higher levels of SPARCK efficacy. In addition, we hope to add to the scarce evidence concerning the change mechanisms in parent training interventions and reveal what works for whom, thereby hopefully allowing us to discover some of the ingredients contained in the “black box”.

By investing in a rigorous design process and incorporating methodological elements that are useful for SPARCK innovation, we hope to address some of the methodological shortcomings of the field of EBI and implementation and to design a novel and tailored intervention that is suitable for evaluation in the context of a large RCT in Norwegian frontline services.

## Data Availability

This is a study protocol for intervention development, no data is used in the article

## References

Ansar, N., Nissen Lie, H. A., Zahl-Olsen, R., Bertelsen, T. B., Elliott, R., & Stiegler, J. R. (2022). Efficacy of Emotion-Focused Parenting Programs for Children’s Internalizing and Externalizing Symptoms: A Randomized Clinical Study. Journal of Clinical Child & Adolescent Psychology, 1–17.

Askeland, E., Forgatch, M. S., Apeland, A., Reer, M., & Grønlie, A. A. (2019). Scaling up an empirically supported intervention with long-term outcomes: The nationwide implementation of GenerationPMTO in Norway. Prevention Science, 20(8), 1189–1199.

Barlow, D. H., Nock, M., & Hersen, M. (2009). Single case experimental designs: Strategies for studying behavior for change.

Bernard, K., Dozier, M., Bick, J., Lewis-Morrarty, E., Lindhiem, O., & Carlson, E. (2012). Enhancing attachment organization among maltreated children: Results of a randomized clinical trial. Child development, 83(2), 623–636.

Braun, V., & Clarke, V. (2006). Using thematic analysis in psychology. Qualitative research in psychology, 3(2), 77–101.

Bølstad, E., Havighurst, S. S., Tamnes, C. K., Nygaard, E., Bjørk, R. F., Stavrinou, M., & Espeseth, T. (2021). A pilot study of a parent emotion socialization intervention: Impact on parent behavior, child self-regulation, and adjustment. Frontiers in psychology, 4552.

Caspi, A., Houts, R. M., Ambler, A., Danese, A., Elliott, M. L., Hariri, A., … Ramrakha, S. (2020). Longitudinal assessment of mental health disorders and comorbidities across 4 decades among participants in the Dunedin birth cohort study. JAMA network open, 3(4), e203221–e203221.

Caspi, A., Houts, R. M., Belsky, D. W., Harrington, H., Hogan, S., Ramrakha, S., … Moffitt, T. E. (2016). Childhood forecasting of a small segment of the population with large economic burden. Nature Human Behaviour, 1, 0005. doi:10.1038/s41562-016-0005

Chen, E., Neta, G., & Roberts, M. C. (2021). Complementary approaches to problem solving in healthcare and public health: implementation science and human-centered design. Translational behavioral medicine, 11(5), 1115–1121.

Child, C. o. t. D. (2022). IDEAS Impact Framework Guiding Principles. Retrieved from: https://developingchild.harvard.edu/innovation-application/innovation-approach/guiding-principles/

Chorpita, B. F., Yim, L., Moffitt, C., Umemoto, L. A., & Francis, S. E. (2000). Assessment of symptoms of DSM-IV anxiety and depression in children: A revised child anxiety and depression scale. Behaviour research and therapy, 38(8), 835–855.

Christiansen, Ø. (2015). Hjelpetiltak i barnevernet-en kunnskapsstatus. [Intervention in child protection services – a reivew]. Retrived from: https://www.bufdir.no/bibliotek/dokumentside/?docid=buf00003222

Collins, L. M. (2018). Optimization of Behavioral, Biobehavioral, and Biomedical Interventions: The Multiphase Optimization Strategy (MOST): Springer.

Comer, J. S., Hong, N., Poznanski, B., Silva, K., & Wilson, M. (2019). Evidence base update on the treatment of early childhood anxiety and related problems. Journal of Clinical Child & Adolescent Psychology, 48(1), 1–15.

Creswell, J. W., & Creswell, J. D. (2017). Research design: Qualitative, quantitative, and mixed methods approaches: Sage publications.

Cyr, C., & Alink, L. R. (2017). Child maltreatment: The central roles of parenting capacities and attachment. Current opinion in psychology, 15, 81–86.

Cyr, C., Euser, E. M., Bakermans-Kranenburg, M. J., & Van Ijzendoorn, M. H. (2010). Attachment security and disorganization in maltreating and high-risk families: A series of meta-analyses. Development and Psychopathology, 22(1), 87–108.

Duncan, G. J., & Magnuson, K. (2013). Investing in preschool programs. Journal of Economic Perspectives, 27(2), 109–132.

Edrissi, F., Havighurst, S. S., Aghebati, A., Habibi, M., & Arani, A. M. (2019). A pilot study of the tuning in to kids parenting program in Iran for reducing preschool children’s anxiety. Journal of Child and Family Studies, 28(6), 1695–1702.

Euser, S., Alink, L. R., Stoltenborgh, M., Bakermans-Kranenburg, M. J., & van IJzendoorn, M. H. (2015). A gloomy picture: a meta-analysis of randomized controlled trials reveals disappointing effectiveness of programs aiming at preventing child maltreatment. BMC public health, 15(1), 1068.

Eyberg, S. M., & Ross, A. W. (1978). Assessment of child behavior problems: The validation of a new inventory. Journal of Clinical Child & Adolescent Psychology, 7(2), 113–116.

Forgatch, M. S., & Kjøbli, J. (2016). Parent Management Training—Oregon Model: Adapting Intervention with Rigorous Research. Family Process.

Forgatch, M. S., & Patterson, G. R. (2010). Parent Management Training—Oregon Model: An intervention for antisocial behavior in children and adolescents. Evidence-based psychotherapies for children and adolescents, 2, 159-178.

Fraire, M. G., & Ollendick, T. H. (2013). Anxiety and oppositional defiant disorder: A transdiagnostic conceptualization. Clinical psychology review, 33(2), 229–240.

Gallo, K. P., Comer, J. S., & Barlow, D. H. (2013). Single-case experimental designs and small pilot trial designs. The Oxford handbook of research strategies for clinical psychology, 24–39.

Auditor general (2021). Riksrevisjonens undersøkelse av psykiske helsetjenester [Auditor General report on mental health services]. (Dokument 3:13). Retrived from: https://www.riksrevisjonen.no/globalassets/rapporter/no-2020-2021/psykiske-helsetjenester.pdf: Governnent

Glasgow, R. E., Vinson, C., Chambers, D., Khoury, M. J., Kaplan, R. M., & Hunter, C. (2012). National Institutes of Health approaches to dissemination and implementation science: current and future directions. American journal of public health, 102(7), 1274–1281.

Goodman, A., Lamping, D. L., & Ploubidis, G. B. (2010). When to use broader internalising and externalising subscales instead of the hypothesised five subscales on the Strengths and Difficulties Questionnaire (SDQ): data from British parents, teachers and children. Journal of abnormal child psychology, 38(8), 1179–1191.

Gottman, J. M., Katz, L. F., & Hooven, C. (1996). Parental meta-emotion philosophy and the emotional life of families: theoretical models and preliminary data. Journal of family psychology, 10(3), 243.

Green, B. L., Ayoub, C., Bartlett, J. D., Furrer, C., Cohen, R. C., Buttita, K., … Sanders, M. B. (2018). How Early Head Start Prevents Child Maltreatment. Retrieved from childtrends.org: https://www.childtrends.org/wp-content/uploads/2018/10/EarlyHeadStartBrief_ChildTrends_October2018.pdf

Greff, M. J., Levine, J. M., Abuzgaia, A. M., Elzagallaai, A. A., Rieder, M. J., & van Uum, S. H. (2018). Hair cortisol analysis: An update on methodological considerations and clinical applications. Clinical biochemistry.

Gubbels, J., van der Put, C. E., & Assink, M. (2019). Risk factors for school absenteeism and dropout: a meta-analytic review. Journal of youth and adolescence, 48(9), 1637–1667.

Johnson, A. M., Hawes, D. J., Eisenberg, N., Kohlhoff, J., & Dudeney, J. (2017). Emotion socialization and child conduct problems: A comprehensive review and meta-analysis. Clinical psychology review, 54, 65-80.

Kazdin, A. E. (2018). Single-case experimental designs. Evaluating interventions in research and clinical practice. Behaviour research and therapy.

Klev, R., & Levin, M. (2016). Forandring som praksis–Endringsledelse gjennom læring og utvikling, 2. utgave, 4. Opplag, Bergen: Fagbokforlaget.

Kornør, H., & Heyerdahl, S. (2017). Måleegenskaper ved den norske versjonen av Strengths and Difficulties Questionnaire, foreldrerapport (SDQ-P).

Kratochwill, T. R., & Levin, J. R. (2014). Enhancing the scientific credibility of single-case intervention research: randomization to the rescue.

Lebowitz, E. R., Panza, K. E., & Bloch, M. H. (2016). Family accommodation in obsessive-compulsive and anxiety disorders: a five-year update. Expert review of neurotherapeutics, 16(1), 45–53.

Lyon, A. R., & Bruns, E. J. (2019). User-centered redesign of evidence-based psychosocial interventions to enhance implementation—hospitable soil or better seeds? JAMA psychiatry, 76(1), 3–4.

Lyon, A. R., Dopp, A. R., Brewer, S. K., Kientz, J. A., & Munson, S. A. (2020). Designing the future of children’s mental health services. Administration and Policy in Mental Health and Mental Health Services Research, 47(5), 735–751.

Lyon, A. R., & Koerner, K. (2016). User-centered design for psychosocial intervention development and implementation. Clinical Psychology: Science and Practice, 23(2), 180–200.

Marchette, L. K., & Weisz, J. R. (2017). Practitioner Review: Empirical evolution of youth psychotherapy toward transdiagnostic approaches. Journal of child psychology and psychiatry, 58(9), 970–984.

Morris, A. S., Criss, M. M., Silk, J. S., & Houltberg, B. J. (2017). The impact of parenting on emotion regulation during childhood and adolescence. Child development perspectives, 11(4), 233–238.

Mummah, S. A., Robinson, T. N., King, A. C., Gardner, C. D., & Sutton, S. (2016). IDEAS (Integrate, Design, Assess, and Share): a framework and toolkit of strategies for the development of more effective digital interventions to change health behavior. Journal of medical Internet research, 18(12).

Nordmo, M., Kinge, J. M., Reme, B.-A., Flatø, M., Surén, P., Wörn, J., … Torvik, F. A. (2022). The educational burden of disease: a cohort study. The Lancet Public Health, 7(6), e549–e556.

Nøkleby, H., Johansen, T. B., Jardim, P. S. J., & Muller, A. E. (2020). Forekomst og behandling av atferdsforstyrrelser: en hurtigoversikt.

Palinkas, L. A., Horwitz, S. M., Chamberlain, P., Hurlburt, M. S., & Landsverk, J. (2011). Mixed-methods designs in mental health services research: a review. Psychiatric Services, 62(3), 255–263.

Patterson, G. R. (2016). Coercion theory: The study of change. The Oxford handbook of coercive relationship dynamics, 7-22.

Proctor, E., Silmere, H., Raghavan, R., Hovmand, P., Aarons, G., Bunger, A., … Hensley, M. (2011). Outcomes for implementation research: conceptual distinctions, measurement challenges, and research agenda. Administration and Policy in Mental Health and Mental Health Services Research, 38(2), 65–76.

Reneflot, A., Aarø, L., Aase, H., Reichborn-Kjennerud, T., Tambs, K., & Øverland, S. (2018). Psykisk helse i Norge Retrieved from fhi.no:

Schindler, H. S., Fisher, P. A., & Shonkoff, J. P. (2017). From innovation to impact at scale: lessons learned from a cluster of research–community partnerships. Child development, 88(5), 1435–1446.

Schindler, H. S., McCoy, D. C., Fisher, P. A., & Shonkoff, J. P. (2019). A historical look at theories of change in early childhood education research. Early childhood research quarterly, 48, 146-154.

Schwartz, C., Barican, J. L., Yung, D., Zheng, Y., & Waddell, C. (2019). Six decades of preventing and treating childhood anxiety disorders: a systematic review and meta-analysis to inform policy and practice. Evidence-based mental health, 22(3), 103–110.

Shimshoni, Y., Shrinivasa, B., Cherian, A. V., & Lebowitz, E. R. (2019). Family accommodation in psychopathology: A synthesized review. Indian journal of psychiatry, 61(Suppl 1), S93.

Shonkoff, J., Richmond, J., Levitt, P., Bunge, S., Cameron, J., Duncan, G., & Nelson III, C. (2016). From best practices to breakthrough impacts a sciencebased approach to building a more promising future for young children and families. Cambirdge, MA: Harvard University, Center on the Developing Child.

Skogen, J., & Torvik, A. (2013). Atferdsforstyrrelser blant barn og unge i Norge. Beregnet forekomst og bruk av hjelpetiltak. [Externalizing problems among Norwegian youth. Prevalence and Services use]. Norwegian Institue of Public Health, report 2013:4.

Smith, J. D. (2012). Single-case experimental designs: a systematic review of published research and current standards. Psychological methods, 17(4), 510.

Solholm, R., Kjøbli, J., & Christiansen, T. (2013). Early Initiatives for Children at Risk—Development of a Program for the Prevention and Treatment of Behavior Problems in Primary Services. Prevention Science, 14(6), 535–544. doi:10.1007/s11121-012-0334-x

Supplee, L. H., & Duggan, A. (2019). Innovative research methods to advance precision in home visiting for more efficient and effective programs. Child development perspectives, 13(3), 173–179.

Technische Universitat Dresden, F. o. P. (2020). Retrieved from https://tu-dresden.de/mn/psychologie/ifap/biopsychologie/forschung/labor/haarsteroidanalyse-und-ptbs-1

Tommeraas, T., & Ogden, T. (2015). Is There a Scale-up Penalty? Testing Behavioral Change in the Scaling up of Parent Management Training in Norway. Administration and Policy in Mental Health and Mental Health Services Research, 1-14. doi:10.1007/s10488-015-0712-3

Waltz, T. J., Powell, B. J., Fernández, M. E., Abadie, B., & Damschroder, L. J. (2019). Choosing implementation strategies to address contextual barriers: diversity in recommendations and future directions. Implementation Science, 14(1), 1–15.

Weisz, J. R., Chorpita, B. F., Frye, A., Ng, M. Y., Lau, N., Bearman, S. K., … Hoagwood, K. E. (2011). Youth top problems: Using idiographic, consumer-guided assessment to identify treatment needs and to track change during psychotherapy. Journal of consulting and clinical psychology, 79(3), 369.

Weisz, J. R., Kuppens, S., Ng, M. Y., Eckshtain, D., Ugueto, A. M., Vaughn-Coaxum, R., … Chu, B. C. (2017). What five decades of research tells us about the effects of youth psychological therapy: a multilevel meta-analysis and implications for science and practice. American Psychologist, 72(2), 79.

Weisz, J. R., Vaughn-Coaxum, R. A., Evans, S. C., Thomassin, K., Hersh, J., Ng, M. Y., … Mair, P. (2019). Efficient monitoring of treatment response during youth psychotherapy: the behavior and feelings survey. Journal of Clinical Child & Adolescent Psychology.

